# Sociodemographic and educational influences on early clinical academic careers in UK medical graduates

**DOI:** 10.64898/2025.12.16.25342392

**Authors:** Emma Fletcher, Katie Petty-Saphon, Hannah Gillespie, Gillian Vance, Lorraine Harper

## Abstract

**Objective:** To examine how sociodemographic and educational factors influence progression into early clinical academic careers among UK medical graduates, focusing on intercalation, Academic Foundation Programme (AFP) participation, and Pre-doctoral Fellowships (PFs).

**Design:** Retrospective cohort study using linked national data from the UK Medical Education Database (UKMED).

**Setting:** All UK medical schools with graduates entering the UK Foundation Programme between 2017 and 2022.

**Participants:** 32,580 graduates (25,290 standard-entry (SE); 7,290 graduate-entry (GE)).

**Main outcome measures:** Completion of an intercalated degree; application to and acceptance for AFP; application to and acceptance for a PF during specialty training.

**Results:** Among SE graduates, 63.9% intercalated, 21.4% applied to AFP and 7.6% were accepted. Male, minority ethnic, privately educated, and less deprived graduates were more likely to intercalate; disability and higher deprivation were associated with lower intercalation. Intercalation, publications, high Education Performance Measure (EPM) scores and attendance at six-year programmes strongly predicted AFP application and acceptance. At PF stage, minority ethnic SE doctors were significantly less likely to be accepted (OR 0.60, 95% CI 0.45 to 0.80, *p* < 0.001), despite similar application rates. AFP completion was the strongest predictor of PF progression (OR 7.82, 95% CI 6.68 to 9.15, *p* <0.001).

GE graduates had similar AFP application (21.9%) and acceptance (6.3%) rates. Male, minority ethnic graduates were more likely to apply to the AFP, while low EPM scores reduced acceptance. Few sociodemographic factors predicted PF applications, although prior AFP acceptance strongly did. Interaction analyses showed GE status attenuated some disparities in AFP application, but reversed patterns observed for ethnicity in acceptance.

**Conclusions:** Early clinical academic progression is shaped by educational achievements and persistent sociodemographic inequalities. Equitable access to funded research opportunities during medical school is likely to strengthen and diversify the future clinical academic workforce.

## INTRODUCTION

Effective translation of scientific discoveries into clinical practice significantly improves patient care. Clinical academics, by combining clinical insight with research expertise, are uniquely positioned to identify gaps in evidence and formulate new research questions, thereby accelerating the transition from basic research to clinical application [1,2]. Hospitals actively engaged in research consistently deliver higher-quality care and report lower mortality rates [3]. However, the clinical academic workforce is in decline, with representation among NHS consultants falling from 4.1% in 2012 to 3.4% in 2024, and the proportion aged over 55 increasing from 17.4% in 2005 to 35.1% in 2024 [4,5]. Maintaining this pipeline is essential as the NHS 10-year plan prioritises innovation and global leadership in research [6]. Additionally, despite the diversity of the UK medical workforce, clinical academics remain predominantly male (64.6%) and white (64.1%), with barriers persisting for women and representation concentrated in research-intensive universities such as Oxford, Cambridge, and Imperial [5–8].

The UK clinical academic pathway is structured as a pipeline beginning with early research engagement during medical school, often through intercalation, followed by entry into the Academic Foundation Programme (AFP; reformed into the Specialised Foundation Programme in 2023) and later pre-doctoral fellowships (PFs), including NIHR Academic Clinical Fellowships (Figure 1). Early exposure to research during undergraduate education, through intercalation and interactions with clinical researchers, has been shown to influence decisions to pursue a clinical academic career [9,10]. However, recent structural changes, specifically the transition from a points-based system to Preference-Informed Allocation for Foundation Training placements, may disincentivize students to intercalate, as the previous system awarded points that improved their ranking for allocation [11]. Additionally, graduate-entry students, who already hold degrees with varying levels of research exposure, previously received extra degree points, but whether they follow an academic trajectory remains unclear. Furthermore, national data on how sociodemographic and educational factors influence progression through this pipeline are lacking. While one study explored extracurricular achievements and demographics [12], comprehensive analysis of socioeconomic background, university type, and academic performance is absent. This is a critical gap given policy initiatives like the NHS Long-Term Workforce Plan aimed at addressing workforce shortages, improving diversity, and expanding medical school places in under-served communities [13].

**Figure 1.**
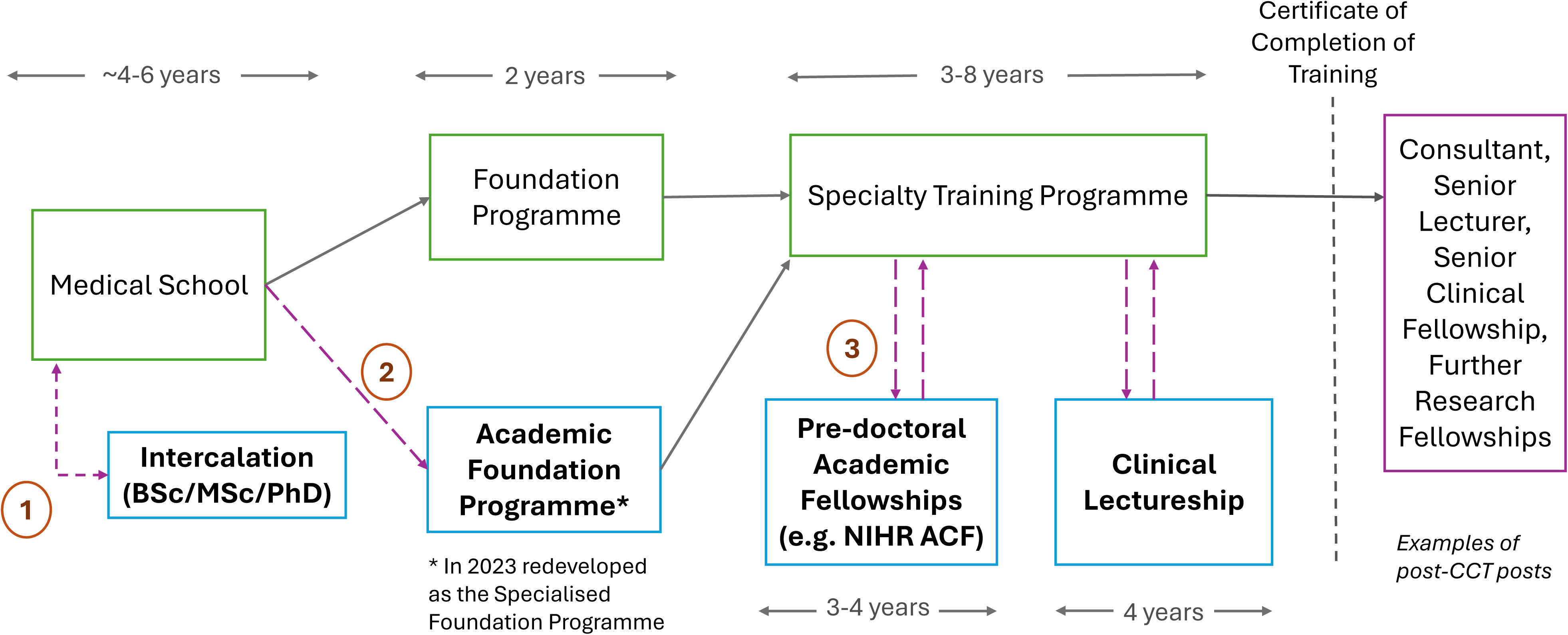
Medical clinical academic training pathway. Key decision points within the structured academic pathway have been labelled (1–3 in orange). These include intercalation, applying to the AFP and applying for a pre-doctoral academic fellowship post, respectively.

This study aims to investigate engagement with intercalation and recruitment to AFPs and PFs using data from the UK Medical Education Database (UKMED). The focus is on sociodemographic and educational factors, with the objective of identifying key predictors of progression and success within the UK clinical academic pathway. We hypothesise that early research exposure, university type, and socioeconomic background significantly influence progression into clinical academia.

## METHODS

### Study design

This is a retrospective cohort study using data from the UKMED, a national database linking undergraduate and postgraduate data to track the progression and outcomes of UK medical students and resident doctors [14]. The study aimed to evaluate whether any sociodemographic and educational factors were associated with progression along the UK clinical academic pathway.

### Data source and setting

Pseudonymised data were accessed after a successful application to UKMED and made available securely through the HIC Dundee Safe Haven Trusted Research Environment. Data were provided for UK-domiciled resident doctors who graduated from a UK medical school between 2017 and 2022 and who directly applied to the UK Foundation Programme.

### Participants

Two cohorts were identified (see Figure 2 for flow diagram): a Standard Entry (SE) cohort, comprising of those who entered medical school as their first degree, and a GE cohort, consisting of those who had previously completed a prior degree. The GE cohort included graduates from GE medicine courses, which are typically completed in four years rather than the standard five to six years for SE courses. It also included individuals who completed a Standard Entry Medicine course as a second degree.

**Figure 2.**
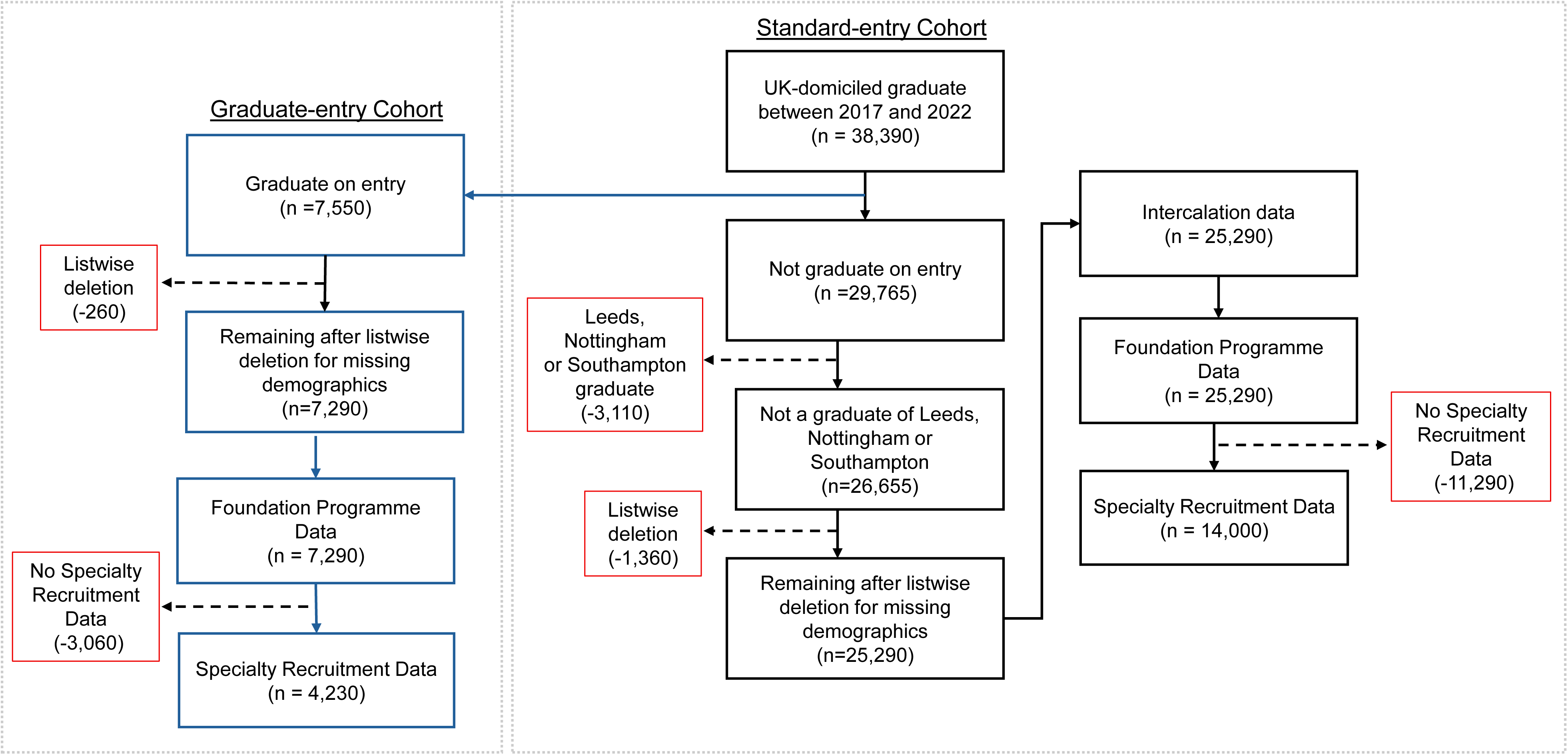
Data flow of the study. Data availability for each outcome and for each Cohort are shown. Cases removed from the study are indicated in red.

For the SE cohort, graduates from Leeds, Nottingham, and Southampton were excluded because their 5-year programmes include a BMedSci, making it difficult to distinguish between mandatory research placements (part of the core curriculum) and additional intercalation. After removing cases with missing sociodemographic or educational data (SE: 1,360; GE: 260) and missing outcome data (SE: 14,350; GE: 3,060), 25,290 SE graduates and 7,290 GE graduates remained for pre-specialty analyses, and 14,000 SE and 4,230 GE graduates for specialty analyses. PF acceptance was not assessed for the GE cohort due to insufficient data.

### Predictors

Several sociodemographic and educational factors were included, though fewer variables were available for the GE cohort due to differences in data availability.

Sociodemographic data were collected on:

- Gender: Defined as male or female, as available in UKMED and included as a covariate in all regression models; associations by gender were estimated within these models rather than through separate stratified analyses.
- Ethnicity: Grouped as Asian/Asian British, Black/Black British, Mixed, Other, and White. For the GE analysis and SE specialty subset, ethnicity was dichotomised into ‘White’ and minority ethnic categories to ensure sufficient statistical power for comparison.
- Index of Multiple Deprivation (IMD): A postcode-based measure collected at entry to medical school. This was dichotomised into ‘most deprived’ (Quintiles 1 and 2) and ‘least deprived’ (Quintiles 3 to 5). This was only included in the SE cohort, as the postcode on entry for GE students may not reliably reflect childhood residence.
- Secondary school type: Grouped by whether students received state- or privately funded education. This was used in the SE cohort only, due to >25% missing data across the GE cohort.
- Disability: Dichotomised into ‘Declared disability’ and ‘None declared’. Used in both cohorts.

Educational factors were defined as:

- Medical School Course Length: Categorised as five-year or six-year courses (where all students intercalate). The six-year schools included Imperial College London, University of Oxford, University of Cambridge, University College London, University of Edinburgh, Queen Mary University of London, and King’s College London.
- Education Performance Measure (EPM): Comprised of three components, each analysed separately:

`o` Medical School Performance: Students were ranked within their graduating cohort based on selected assessments. Graduates were grouped into ‘low’ (deciles 6–10) and ‘high’ (deciles 1–5) performance categories.
`o` Intercalation: Presence of intercalation was inferred from awarded points. This was only used in the SE cohort, as prior degrees in the GE cohort were likely from their primary degree.
`o` · Publication: Graduates could receive points for publishing a peer-reviewed journal paper. Presence of points indicated a publication. Used only in the SE cohort, as it was difficult to distinguish potential publications from the primary degree in the GE cohort.
`o` · Medical School Type: Dichotomised into Traditional (pre-1992 accreditation) or Modern (post-1992 accreditation). Traditional medical schools are often more research-intensive and emphasise deeper standalone theoretical science with a pre-clinical/clinical divide, while modern schools use a more integrated approach.

### Outcomes

Although the study focused on the early clinical academic pipeline, individual milestones were assessed separately. The primary outcomes included whether graduates had intercalated during medical school, applied for an AFP, and were accepted to AFP. It further included whether graduates applied for or were accepted to a pre-doctoral fellowship (PF) during specialty training.

### Bias

Selection bias was minimised by including all eligible graduates with complete data. Missing data were decidedly handled using listwise deletion. Although complete-case analysis slightly reduced the sample size, the loss did not materially affect study power or estimates.

### Sample size

A priori power analysis using G*Power [15] indicated that detecting an odds ratio of 1.2 with 80% power and a one-sided α of 0.01, required 1808 participants. All cohort groups exceeded this minimum, ensuring sufficient statistical power for the analyses.

### Statistical analysis

Analyses were performed in R Studio (version 4.4.0) within the Dundee HIC TRE Safe Haven. Descriptive statistics and a Chi-squared Test of Independence (*χ²*) were initially used to identify univariate associations between characteristics and each cohort or academic pathway outcome. These are presented in Supplementary Tables 1 and 2.

Separate multivariate analyses were conducted using logistic regression models for the SE and GE cohorts to assess sociodemographic and educational variables. This cohort-specific approach allowed for tailored analysis, accounting for differences in sociodemographic and outcome availability. We also fitted a combined model with interaction terms for course type (SE vs GE) to assess moderation effects. This interaction model provided a more nuanced understanding of how course type may moderate the relationships between sociodemographic factors and early clinical academic career outcomes.

Additionally, for the intercalation model, only demographic variables were included due to the temporal ordering of educational factors. Cramer’s V was used to assess the strength of associations between sociodemographic and educational variables, helping to minimise multicollinearity (Supplementary Figures 1 and 2). Sensitivity analyses were conducted by varying model specifications, including alternative combinations of sociodemographic and educational variables, to test the robustness of results. Additional post-hoc evaluations were conducted, including goodness-of-fit testing, chi-square tests to assess associations between variables, ROC analysis to evaluate discriminatory performance, and residual analysis to assess model fit. These analyses indicated that the models were appropriately specified and robust. *P*-values below 0.05 were considered to indicate statistical significance.

### Ethics

The project was approved by the Science, Technology, Engineering and Mathematics Research Committee at the University of Birmingham (reference no 2819).

### Patient and Public Involvement

No patients or members of the public were involved in the study.

## RESULTS

Outcome data for intercalation and application to the UKFP were available for both full cohorts (N = 25,290 SE and N = 7,290 GE). A smaller subset (N = 14,000 SE and N = 4,230 GE) had data available for application to specialty training. Demographic, socioeconomic, and educational characteristics are summarised in Table 1. The specialty subgroups were broadly representative of their respective cohorts

**Table 1.**
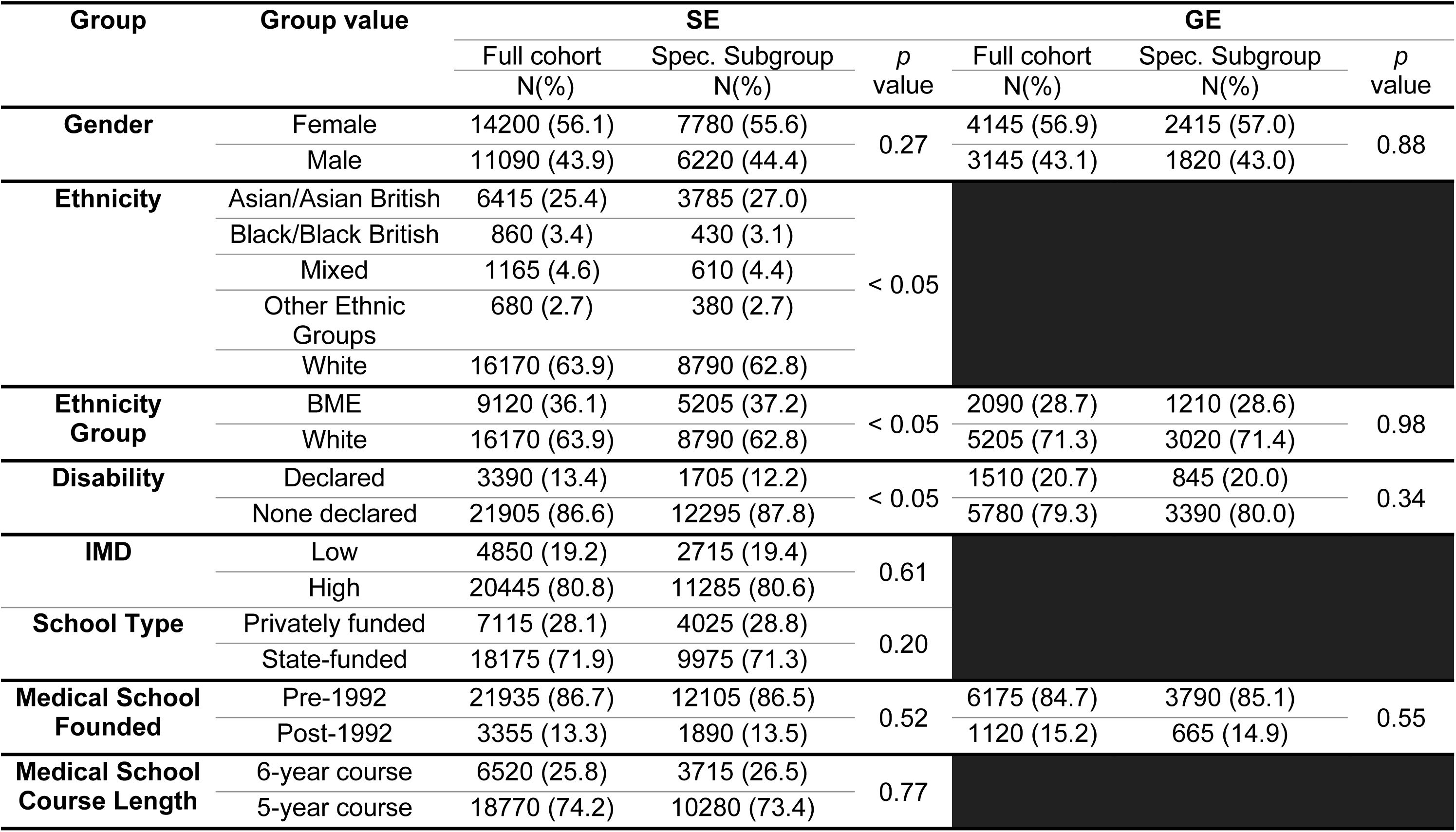
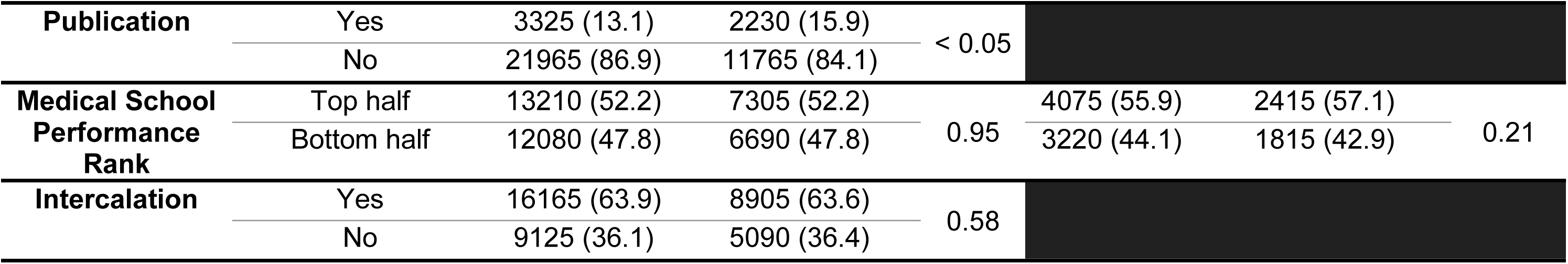
Demographic characteristics of the standard-entry (SE) and graduate-entry (GE) full cohorts and specialty (spec.) subgroups. Some demographic and educational variables were not available or applicable for the GE cohort. Chi-squared (χ²) tests were used to compare proportions between the full cohorts and their respective specialty subgroups to ensure subgroup comparability.

| Group | Group value | SE |  |  | GE |  |  |
| --- | --- | --- | --- | --- | --- | --- | --- |
|  |  | Full cohort<br>N(%) | Spec. Subgroup<br>N(%) | *p*<br>value | Full cohort<br>N(%) | Spec. Subgroup<br>N(%) | *p*<br>value |
| Gender | Female | 14200 (56.1) | 7780 (55.6) | 0.27 | 4145 (56.9) | 2415 (57.0) | 0.88 |
|  | Male | 11090 (43.9) | 6220 (44.4) |  | 3145 (43.1) | 1820 (43.0) |  |
| Ethnicity | Asian/Asian British | 6415 (25.4) | 3785 (27.0) | < 0.05 |  |  |  |
|  | Black/Black British | 860 (3.4) | 430 (3.1) |  |  |  |  |
|  | Mixed | 1165 (4.6) | 610 (4.4) |  |  |  |  |
|  | Other Ethnic Groups | 680 (2.7) | 380 (2.7) |  |  |  |  |
|  | White | 16170 (63.9) | 8790 (62.8) |  |  |  |  |
| Ethnicity Group | BME | 9120 (36.1) | 5205 (37.2) | < 0.05 | 2090 (28.7) | 1210 (28.6) | 0.98 |
|  | White | 16170 (63.9) | 8790 (62.8) |  | 5205 (71.3) | 3020 (71.4) |  |
| Disability | Declared | 3390 (13.4) | 1705 (12.2) | < 0.05 | 1510 (20.7) | 845 (20.0) | 0.34 |
|  | None declared | 21905 (86.6) | 12295 (87.8) |  | 5780 (79.3) | 3390 (80.0) |  |
| IMD | Low | 4850 (19.2) | 2715 (19.4) | 0.61 |  |  |  |
|  | High | 20445 (80.8) | 11285 (80.6) |  |  |  |  |
| School Type | Privately funded | 7115 (28.1) | 4025 (28.8) | 0.20 |  |  |  |
|  | State-funded | 18175 (71.9) | 9975 (71.3) |  |  |  |  |
| Medical School Founded | Pre-1992 | 21935 (86.7) | 12105 (86.5) | 0.52 | 6175 (84.7) | 3790 (85.1) | 0.55 |
|  | Post-1992 | 3355 (13.3) | 1890 (13.5) |  | 1120 (15.2) | 665 (14.9) |  |
| Medical School Course Length | 6-year course | 6520 (25.8) | 3715 (26.5) | 0.77 |  |  |  |
|  | 5-year course | 18770 (74.2) | 10280 (73.4) |  |  |  |  |
| Publication | Yes | 3325 (13.1) | 2230 (15.9) | < 0.05 |  |  |  |
|  | No | 21965 (86.9) | 11765 (84.1) |  |  |  |  |
| Medical School<br>Performance<br>Rank | Top half | 13210 (52.2) | 7305 (52.2) | 0.95 | 4075 (55.9) | 2415 (57.1) | 0.21 |
|  | Bottom half | 12080 (47.8) | 6690 (47.8) |  | 3220 (44.1) | 1815 (42.9) |  |
| Intercalation | Yes | 16165 (63.9) | 8905 (63.6) | 0.58 |  |  |  |
|  | No | 9125 (36.1) | 5090 (36.4) |  |  |  |  |

**Table 2.**
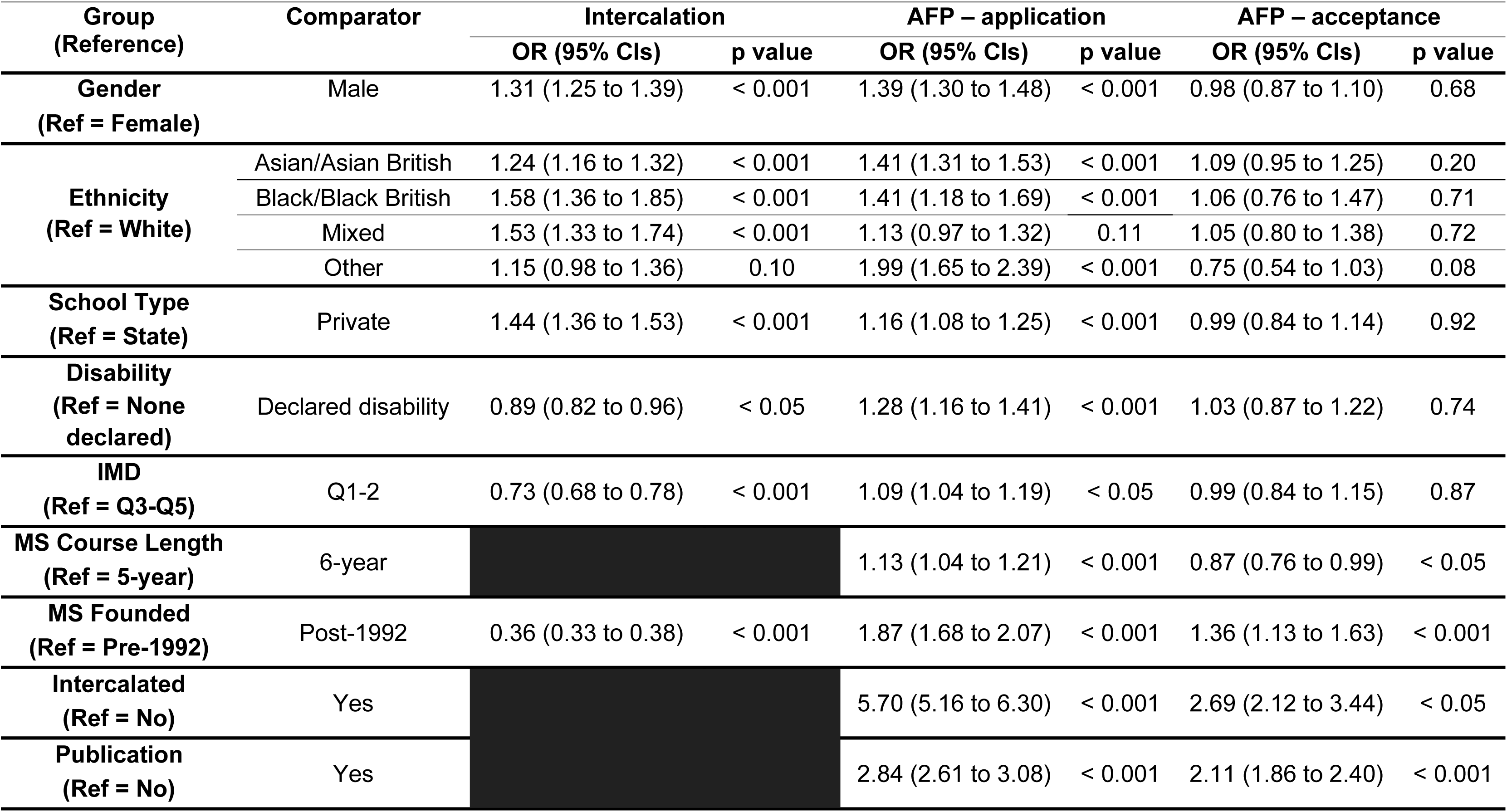
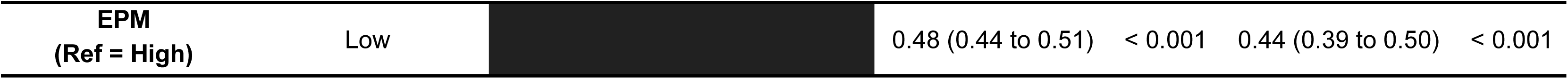
Multivariate analysis of sociodemographic and educational factors associated with intercalating, applying to, and accepting an Academic Foundation Programme (AFP) offer in the standard-entry cohort. .

### Sociodemographic differences between SE and GE cohorts

Across the shared sociodemographic variables, there were some differences between the SE and GE cohorts. Gender distribution was comparable (SE: 56.1% female, GE: 56.9%), as was the proportion of students from pre-1992 medical schools (SE: 86.7%, GE: 84.7%). The GE cohort was less ethnically diverse, with 28.7% from an ethnic minority background compared to 36.1% in the SE cohort. A higher proportion of GE students reported a declared disability (28.7% vs. 13.4% SE). GE graduates were also slightly more likely to rank in the top half of medical school performance (55.9% compared to 52.2% in the SE cohort) and to be from a modern medical school (GE: 15.2%, SE: 13.3%).

### Impact of sociodemographic factors on the SE cohort on early clinical academic careers

In total 63.9% of the SE cohort intercalated, stable across 5-years. In the multivariate analysis (Table 2), male graduates were more likely to have intercalated than female (OR 1.31, 95% CI 1.25 to 1.39, *p* < 0.001). All ethnic minority groups had an increased odds of intercalation compared to White ethnicity, with Black ethnicity showing the highest likelihood of intercalating (OR 1.58, 95% CI 1.36 to 1.85, *p* < 0.001). Individuals who attended a private school were more likely to have intercalated than those from state schools (OR 1.44, 95% CI 1.36 to 1.53, *p* < 0.001), while those from more deprived IMD quintiles (OR 0.73, 95% CI 0.68 to 0.78, *p* < 0.001) or who had a declared a disability (OR 0.89, 95% CI 0.68 to 0.78, *p* < 0.001) has reduced odds.

In total, 21.4% of graduates applied and 7.6% were accepted on to an AFP. As seen for intercalation, male graduates, most ethnic minority groups, except those identifying as Mixed, and those who had attended a private school were more likely to apply for AFP. However, graduates from the most deprived IMD quintiles (OR 1.09, 95% CI 1.04 to 1.19, *p* < 0.05) and those declaring a disability (OR 1.28, 95% CI 1.16 to 1.41, *p* < 0.001) were more likely to apply. No significant variations in acceptance rates were observed within demographic groups.

Among the 14,000 SE doctors with specialty data, 7.5% applied and 2.8% were accepted on to PF programmes. Male doctors were more likely to apply than female doctors (OR 1.23, 95% CI 1.07 to 1.41, *p* < 0.001), though acceptance rates did not differ. No significant difference in application rates was observed between minority ethnic and White doctors, although minority ethnic doctors were far less likely to be accepted (OR 0.60, 95% CI 0.45 to 0.80, *p* < 0.001). There was no difference for IMD quintiles or disability (Table 3).

**Table 3.** Multivariate analysis of sociodemographic and educational factors associated with applying to and accepting a pre-doctoral fellowship (PF) in the standard-entry cohort. .

| Group<br>(Reference) | Comparator | PF – application |  | PF – acceptance |  |
| --- | --- | --- | --- | --- | --- |
|  |  | OR (95% CIs) | p value | OR (95% CIs) | p value |
| **Gender**<br>(Ref = Female) | Male | 1.23 (1.07 to 1.41) | < 0.05 | 1.00 (0.76 to 1.30) | 0.99 |
| **Ethnicity Group**<br>(Ref = White) | Ethnic minority | 1.08 (0.93 to 1.25) | 0.31 | 0.60 (0.45 to 0.80) | < 0.001 |
| **School Type**<br>(Ref = State) | Private | 1.08 (0.93 to 1.25) | 0.31 | 1.48 (1.12 to 1.97) | < 0.05 |
| **Disability**<br>(Ref = None declared) | Declared disability | 1.11 (0.90 to 1.36) | 0.33 | 1.11 (0.73 to 1.65) | 0.64 |
| **IMD**<br>(Ref = Q3-Q5) | Q1-2 | 0.96 (0.80 to 1.16) | 0.70 | 0.93 (0.63 to 1.34) | 0.68 |
| **6-year MS course**<br>(Ref = No) | Yes | 1.20 (1.03 to 1.40) | < 0.05 | 1.01 (0.75 to 1.36) | 0.94 |
| **MS Founded**<br>(Ref = Pre-1992) | Post-1992 | 1.11 (0.88 to 1.39) | 0.38 | 1.35 (0.87 to 2.09) | 0.18 |
| **Intercalated**<br>(Ref = No) | Yes | 3.18 (2.54 to 4.01) | < 0.001 | 1.35 (0.83 to 2.46) | 0.22 |
| **Publication**<br>(Ref = No) | Yes | 1.91 (1.64 to 2.23) | < 0.001 | 1.33 (1.00 to 1.78) | 0.05 |
| **EPM**<br>(Ref = High) | Low | 0.88 (0.76 to 1.03) | 0.11 | 0.63 (0.46 to 0.85) | < 0.05 |
| **FP type**<br>(Ref = FP) | AFP | 7.82 (6.68 to 9.15) | < 0.001 | 2.86 (2.16 to 3.80) | < 0.001 |

### Impact of educational factors on the SE cohort on early clinical academic careers

Intercalation (OR 5.70, 95% CI 5.16 to 6.30, *p* < 0.001) and peer-reviewed publications (OR 2.84, 95% CI 2.61 to 3.08, *p* < 0.001) were strongly associated with increased odds of application to AFP, as well as acceptance (OR 2.69, 95% CI 2.12 to 3.44, *p* < 0.001 and OR 2.11, 95% CI 1.86 to 2.40, *p* < 0.001). Low EPM scores were found to reduce both application (OR 0.47, 95% CI 0.44 to 0.51, *p* < 0.001) and acceptance for AFP (OR 0.48, 95% CI 0.44 to 0.51, *p* < 0.001). Graduates from 6-year programmes had higher AFP application rates (OR 1.13, 95% CI 1.04 to 1.21, *p* < 0.001) but were less likely to be accepted (OR 0.87, 95% CI 0.76 to 0.99, *p* < 0.05). While graduates from a modern medical school were far less likely to intercalate (OR 0.36, CI 0.33 to 0.38, *p* < 0.001), they were more likely to apply to AFP and be accepted (OR 1.87, CI 1.68 to 2.07, *p* < 0.001 and OR 1.36, 95% CI 1.13 to 1.63, *p* < 0.001).

Educational factors showed stronger associations for PF application and success than sociodemographic factors. Intercalation (OR 3.18, 95% CI 2.54 to 4.01, *p* < 0.001) and 6-year medical courses (OR 1.20, 95% CI 1.03 to 1.40, *p* < 0.05) were linked to higher odds of application, but not acceptance. Publications were linked to both a higher odds of application (OR 1.91, 95% CI 1.64 to 2.23, *p* < 0.001) and acceptance (OR 1.33, 95% CI 1.00 to 1.78, *p* < 0.05). A low EPM score reduced acceptance likelihood (OR 0.61, 95% CI 0.45 to 0.83, *p* < 0.001). Those who completed the AFP were significantly more likely to apply to (OR 7.82, 95% CI 6.68 to 9.15, *p* < 0.001) and be accepted for (OR 2.86, 95% CI 2.16 to 3.80, *p* < 0.001) a PF than those completing the standard FP. The medical school founding year had no effect.

### Impact of sociodemographic and educational factors on the GE cohort on early clinical academic careers

Among GE graduates, 21.9% of graduates applied to the AFP, with 6.3% successfully accepted. Consistent with the SE cohort, male graduates and those from ethnic minority backgrounds were more likely to apply to the AFP, while those with a low EPM score were less likely (Table 4). Contrasting to the SE cohort, graduates with a declared disability were less likely to apply (OR 0.84, 95% CI 0.74 to 0.97, *p* < 0.05). Significant differences in acceptance rates were observed for male graduates (OR 1.30, 95% CI 1.07 to 1.57, *p* < 0.05) and those with a low EPM score (OR 0.46, 95% CI 0.37 to 0.57, *p* < 0.001). No sociodemographic or educational factors were significantly associated with a PF application, except for prior acceptance to the AFP (OR 9.68, 95% CI 7.18 to 13.02, *p* < 0.001).

**Table 4.**
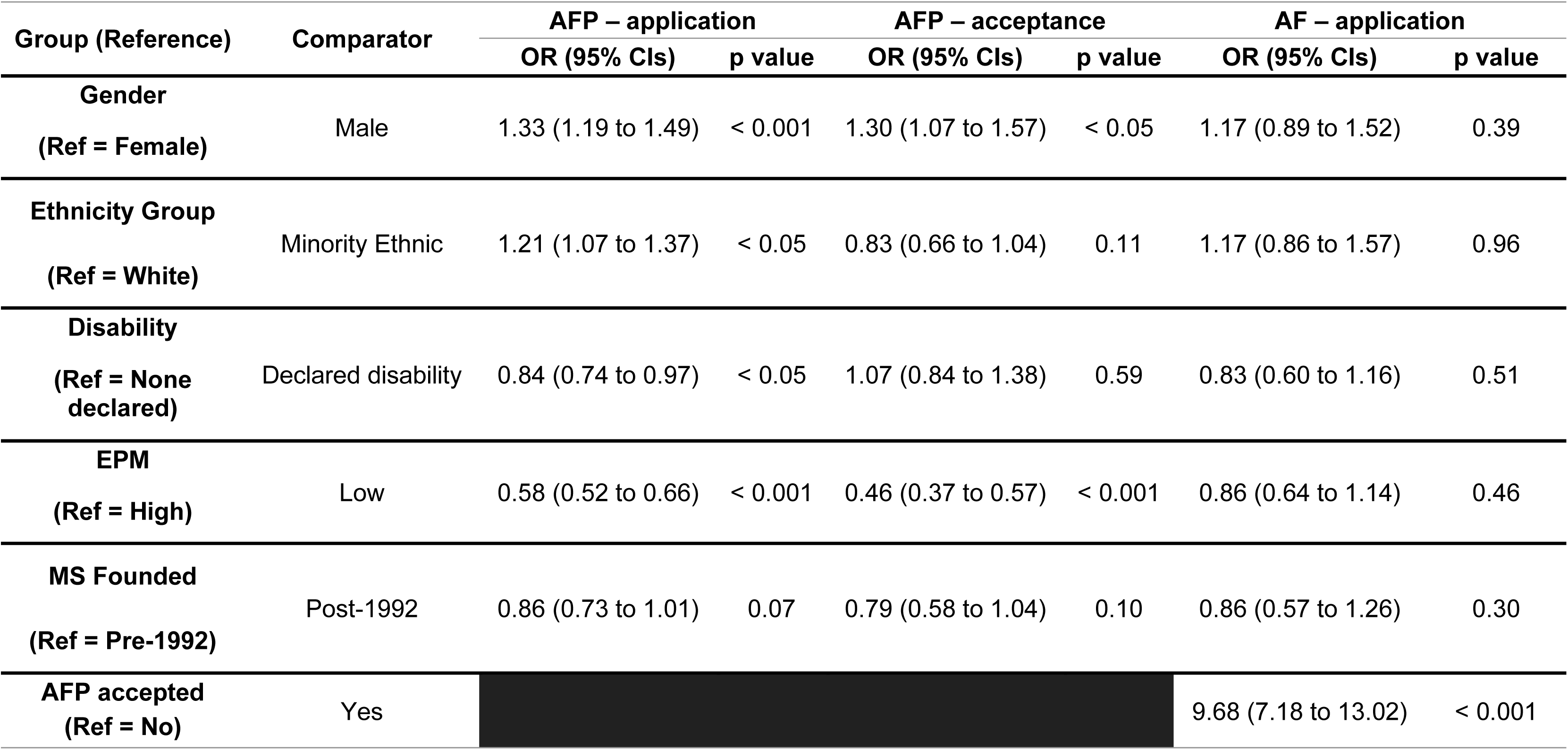
Multivariate analysis for the association of sociodemographic and educational factors with applying to and accepting to the Academic Foundation Programme (AFP), and application to an pre-doctoral fellowship (PF) for the graduate-entry cohort. .

### Impact of graduate entry status on sociodemographic and educational associations

The results of the combined model have been plotted in (Figure 3; Supplementary Tables 3-5). No significant variations were observed between the GE and SE groups for overall rates of application to the AFP or an AF, or acceptance to AFP.

**Figure 3.**
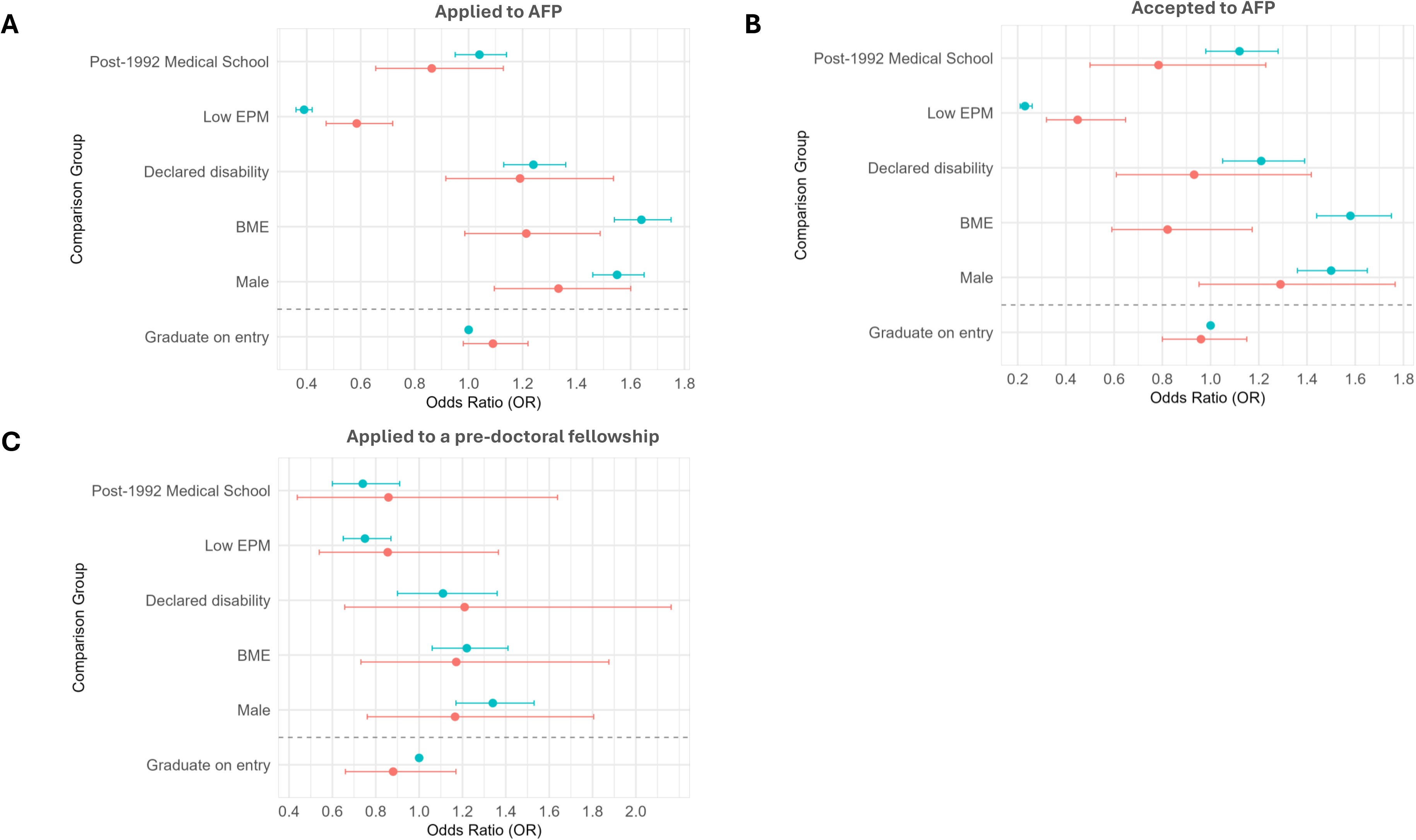
Odds ratios comparing graduate-entry and standard-entry applicants across three stages of the academic pathway. Each subplot represents a distinct stage: (A) Application to the Academic Foundation Programme (AFP), (B) Acceptance to the AFP, and (C) Application to a pre-doctoral fellowship. Odds ratios are presented with 95% confidence intervals. For the ‘graduate on entry’ comparison, standard-entry is treated as the reference group with an odds ratio of 1.00. All other comparisons incorporate interaction effects to reflect differences in odds across demographic characteristics.

For AFP application, the interaction term for GE status attenuated several associations, such as reducing the increased odds for males (interaction OR 0.86, 95% CI 0.75 to 0.97, *p* < 0.05) and those from ethnic minority backgrounds (interaction OR 0.74, 95% CI 0.64 to 0.85, *p* < 0.001), as well as increasing the odds associated with a low EPM score (interaction OR 1.50, 95% CI 1.31 to 1.71, *p* < 0.001). In contrast, GE graduates from post-1992 institutions had significantly lower odds of applying (interaction OR 0.83, 95% CI 0.69 to 0.99, *p* < 0.05). No significant variation was observed for those with a declared disability.

For AFP acceptance, applicants from an ethnic minority background had higher odds of success in the SE group than white applicants. This pattern was reversed with GE graduates (interaction OR 0.52, 95% CI 0.41 to 0.67, *p* < 0.001). The GE status reduced the negative effect of a low EPM score on acceptance (interaction OR 1.95, 95% CI 1.52 to 2.49, *p* < 0.001) and reduced the effect seen for GE graduates from newer medical schools being accepted on to AFP programmes (interaction OR 0.70, 95% CI 0.51 to 0.96, *p* < 0.05). There were no significant differences linked with GE status for those with a declared disability or who were male. For PF applications, GE interactions were not statistically significant, providing no evidence that GE status modified any associations.

## DISCUSSION

This national cohort study of over 32,000 UK medical graduates provides novel insights into the educational and sociodemographic factors that shape progression into early clinical academic careers. By examining intercalation, applications to the AFP, and subsequent PFs, we demonstrate that both structural and individual factors influence career trajectories in clinical academia. Our findings highlight disparities in access and success at each stage, raising important questions about equity, inclusivity, and the sustainability of the clinical academic workforce.

### Gender

We observed that male graduates were consistently more likely to intercalate, apply for AFP, and apply for PF posts than female graduates, irrespective of graduate status, although acceptance rates for male graduates were increased within the GE cohort. This pattern suggests that the gender imbalance in clinical academia may not result from differences in ability or selection but from earlier decisions by women not to pursue academic pathways. Previous literature has identified barriers including limited female role models, challenges in work–life balance, and perceptions of academic careers as incompatible with family life [16,17]. These challenges may be more pronounced in the older GE cohort, possibly explaining the additional impact on acceptance rates. Early intervention during medical school - through mentorship, visibility of successful female academics, and flexible research opportunities - may be crucial in raising interest [18].

### Ethnicity

Ethnic minority graduates were more likely to intercalate and both GE and SE cohorts were more likely to apply for AFP, suggesting strong interest in academic opportunities. However, striking disparities were seen within the SE cohort at the PF stage, where minority ethnic doctors were significantly less likely to be accepted despite similar application rates. This raises concerns about structural barriers, potential bias in selection processes, and inequities in access to mentoring or networks that support progression [19,20]. Efforts to ensure transparency in recruitment, alongside targeted mentorship and support for applicants from a minority ethnic background, are needed to address this gap [21].

### School type and socioeconomic background (SE only)

Graduates from private schools were more likely to intercalate and apply to AFP, possibly reflecting the advantages of prior educational opportunities [12]. Despite similar acceptance rates at the AFP stage, private school background appeared to confer an advantage in PF acceptance, suggesting that early educational privilege may translate into later academic opportunities.

Socioeconomic background, as measured by IMD, presented a more complex picture. Graduates from more deprived backgrounds were less likely to intercalate but paradoxically more likely to apply for AFP, with no observed differences in acceptance. This may reflect the added cost of an intercalating year but awareness of the career-enhancing potential of AFP posts among less advantaged groups [10]. Widening participation initiatives in medical schools have been shown to improve diversity but need further embedding to ensure equitable access to funded intercalated degrees and research experiences [22,23].

### Disability

Graduates who declared a disability were less likely to have intercalated and both cohorts were more likely to apply for AFP, yet acceptance rates were equivalent to those without a declared disability. No differences were seen at the PF stage. These findings may reflect growing awareness and support for disability inclusion in early postgraduate training [24]. Nonetheless, more qualitative research is required to explore motivations for applying, experiences of disabled doctors within academic training, and whether structural barriers persist at later career stages [25,26].

### Educational factors

Educational markers had stronger associations with academic progression than sociodemographic factors. Intercalation and peer-reviewed publications during medical school were robust predictors of both AFP application and acceptance, reinforcing their value as early indicators of academic engagement [27,28]. These achievements may further reflect access to formal mentorship and a supportive environment, which subsequently has a positive influence of career trajectory, through increased confidence, research skills and academic support. This study provides added value given the national nature of the study as most previous studies are based on small cohorts. While intercalation and peer-reviewed publications also predicted PF applications, only peer-reviewed publications were associated with acceptance suggesting that successful engagement and outputs foster success irrespective of where the research experience comes from. However, this may also reflect enhanced mentorship and support which is not equitably available. Bain et al. found that students without prior research experience were significantly more likely to note that a lack of research exposure and limited understanding of academic career pathways as key barriers [29]. This suggests that early engagement in research both enhances confidence and fosters greater awareness of academic career opportunities.

The strength of the clinical academic pipeline relies on early and equitable access to research opportunities. Until 2023, the EPM encouraged academic engagement by awarding UKFP points for achievements such as publications and intercalated degrees [11]. The introduction of the Preference Informed Allocation (PIA) system, which allocates posts based on ranked preferences and random selection, in 2023 replaced this points-based approach to improve fairness and reduce stress. However, has been reported to disincentivise medical students from pursuing research during their degree, as it no longer directly impacts on their chances of securing preferred posts. [10]. Ensuring that all medical students can access meaningful research opportunities remains essential.

SE graduates from six-year courses were more likely to apply for PFs but showed no variance in acceptance rate. Published literature has suggested that mandatory intercalation fosters interest in academia [29] and there is a greater proportion of graduates from universities with mandatory intercalation in academic training posts [8], yet our data show it does not confer an advantage over traditional 5-year courses.

Low EPM scores had the same effect for both SE and GE cohorts in that they reduced both AFP applications and acceptance, consistent with their role in ranking applicants [30]. At the PF level, low EPM scores only influenced acceptance, suggesting that once applicants self-select into the pathway, academic performance remains an important determinant of success.

Finally, completion of the AFP strongly predicted both application to and acceptance into an PF, underlining the AFP as a critical stepping stone for future clinical academics. This finding highlights the importance of maintaining the integrity and visibility of AFP posts within postgraduate training [31].

### Graduate-entry status

Direct comparisons showed no overall significant differences in application or acceptance rates between SE and GE graduates. This is reassuring given previous studies have shown no difference or better outcomes in performance between these two cohorts during the clinical phase of the undergraduate course [32–36]. However, GE status influenced several demographic disparities in AFP outcomes. GE graduates had lowered odds of acceptance for males and ethnic minority applicants compared to SE, while GE graduates from post-1992 institutions had lower odds of acceptance than SE graduates. The positive association between ethnic minority status and AFP acceptance observed in SE graduates was reversed for GE graduates, suggesting a possible intersectional relationship. As demonstrated here and by Garrud and McManus [36], GE medicine has a higher proportion of White graduates compared to SE medicine courses, which may translate into a lack of relatable role models for ethnic minority GE students. GE graduates appeared less impacted by low EPM scores than SE graduates, while no significant effects were observed for disability or PF applications.

### Strengths and limitations

The study draws strength from its large, nationally representative cohort, use of robust linked datasets, and examination of multiple stages in the clinical academic pathway. However, limitations include exclusion of some medical schools and graduates with missing data, the simplification of complex sociodemographic variables, and incomplete specialty training data, limiting longer-term outcomes such as clinical lectureships. Additionally, results were not disaggregated by sex or gender. Although the cohort was sufficient, separate analyses were not justified by the study design; instead, gender was included as a covariate in all models to account for its potential influence. Finally, intersectionality between gender, ethnicity, and socioeconomic status was not fully explored but represents an important direction for future research [37].

### Implications

Our findings highlight key disparities in early clinical academic progression for both SE and GE graduates, particularly affecting women, ethnic minority doctors, and those from lower socioeconomical backgrounds. Educational experiences such as intercalation and publications remain strong predictors of success, but inequitable access to these opportunities risks perpetuating inequalities in the clinical academic workforce.

Sustaining the pipeline of clinical academics requires nurturing interest among medical students while also ensuring that progression is based on academic ability and not constrained by background or structural barriers.

There has been an increasing awareness of the threats to clinical academic careers. A recent task-and-finish group commissioned by the OSCHR has issued recommendations to address these threats, including a national scheme to support early research experience through funded intercalated degree bursaries and internships, and selection criteria in foundation training that recognise research experience and motivation; reversing recent policy decisions [4]. The NIHR has already introduced funding for recently accredited medical schools to provide bursaries for intercalation and research experiences. This study supports these recommendations and suggests investment in undergraduate research experience is likely to impact longer term academic career progression.

### What is already known on this topic

- Early research exposure, including intercalated degrees, is thought to influence the pursuit of clinical academic careers. However, no UK-wide analyses have examined how sociodemographic and educational factors affect progression along these pathways.
- Understanding these predictors is important in the context of a declining and ageing clinical academic workforce, ongoing challenges in diversity, and recent policy changes affecting medical education.

### What this study adds

- For standard-entry (SE) graduates, intercalation, publications, high EPM scores, and academic foundation posts most strongly predict progression along the clinical academic pathway, while barriers persist still for women, minority ethnic doctors, and those from deprived backgrounds.
- Graduate-entry status mitigated some sociodemographic disparities, but inequalities remain, particularly for ethnic minority doctors, emphasising the need for equitable access to research opportunities across all entry routes.

### Strengths and limitations of this study

#### Strengths

- Large, nationally representative cohort of over 32,000 UK medical graduates, ensuring robust statistical power.
- Use of linked UKMED datasets enabled tracking across multiple stages of the clinical academic pathway.

#### Limitations

- Exclusion of some medical schools and graduates with missing data may limit generalisability.
- Simplification of complex sociodemographic variables and incomplete specialty training data restricts analysis of longer-term outcomes.
- Intersectionality between gender, ethnicity, and socioeconomic status was not fully explored.

## ETHICAL APPROVAL

The project was approved by the Science, Technology, Engineering and Mathematics Research Committee at the University of Birmingham (reference no 2819). No patients or members of the public were involved in the study.

## DATA AVAILABILITY STATEMENT

The data used in this study were accessed through the HiC Dundee Safe Haven under the UKMED application process. Access was granted following approval of our application and signing of a Data Sharing Agreement (DSA). Data were provided in pseudonymised form and were stored in a secure environment; therefore, they cannot be shared publicly. Researchers who meet the criteria for access can request access via the UKMED application process and use of the HiC Dundee Safe Haven.

The source of the data used in this project is the UK Medical Education Database (UKMED) [project number P206], with the extract generated on 04/05/2025. Approved for publication on 19th June 2025. I am grateful to UKMED for the use of these data. However, UKMED bears no responsibility for their analysis or interpretation. The data includes information derived from that collected by the Higher Education Statistics Agency Limited (“HESA”) and provided to the GMC (“HESA Data”), source: HESA Student Record 21051. Copyright Higher Education Statistics Agency Limited. The Higher Education Statistics Agency Limited makes no warranty as to the accuracy of the HESA Data and cannot accept responsibility for any inferences or conclusions derived by third parties from data or other information supplied by it.

## FOOTNOTES

### Author contributions

EF is the guarantor of the study. EF and LH conceived the study with support from KPS and applied for access to the data. EF performed the statistical analysis. EF and LH interpreted the data with support from HG, GV, and KPS. EF and LH drafted the manuscript. EF, LH, HG, GV, and KPS critically revised the manuscript. All authors approved the final version.

### Transparency statement

The lead author (guarantor) affirms that this manuscript is an honest, accurate, and transparent account of the study being reported; that no important aspects of the study have been omitted; and that any discrepancies from the study as originally planned have been explained.

### Dissemination to participants and related patient and public communities

We plan to present our findings at relevant national and international forums and conferences to reach relevant stakeholders. We also plan to use social media outlets to disseminate findings.

### Provenance and peer review

Not commissioned; externally peer reviewed.

