## Supplementary materials for "Sociodemographic and educational influences on early clinical academic careers in UK medical graduates"

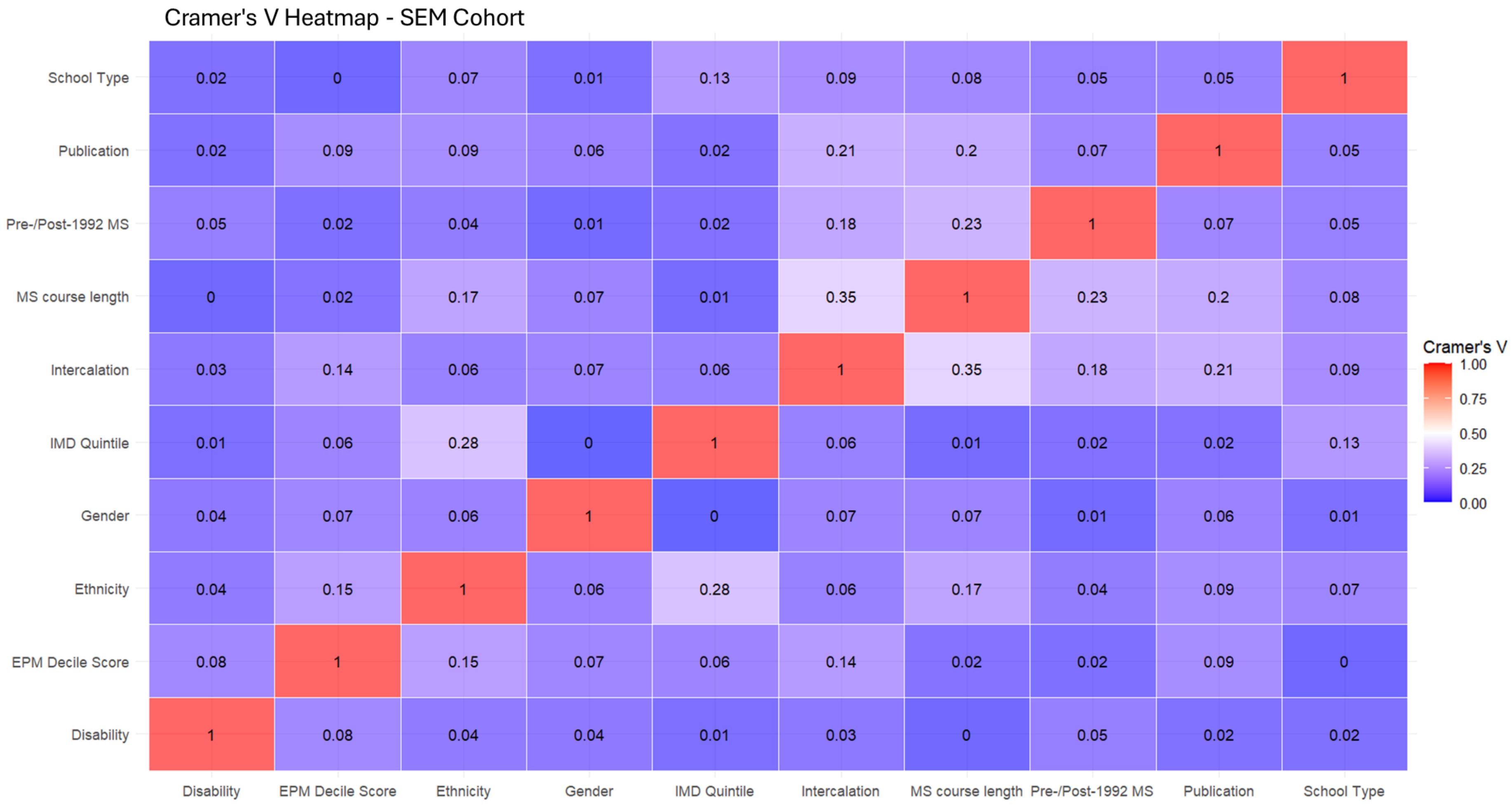


**Supplementary Figure 1: Cramer's V showing the association of demographic variables in the SE cohort.**

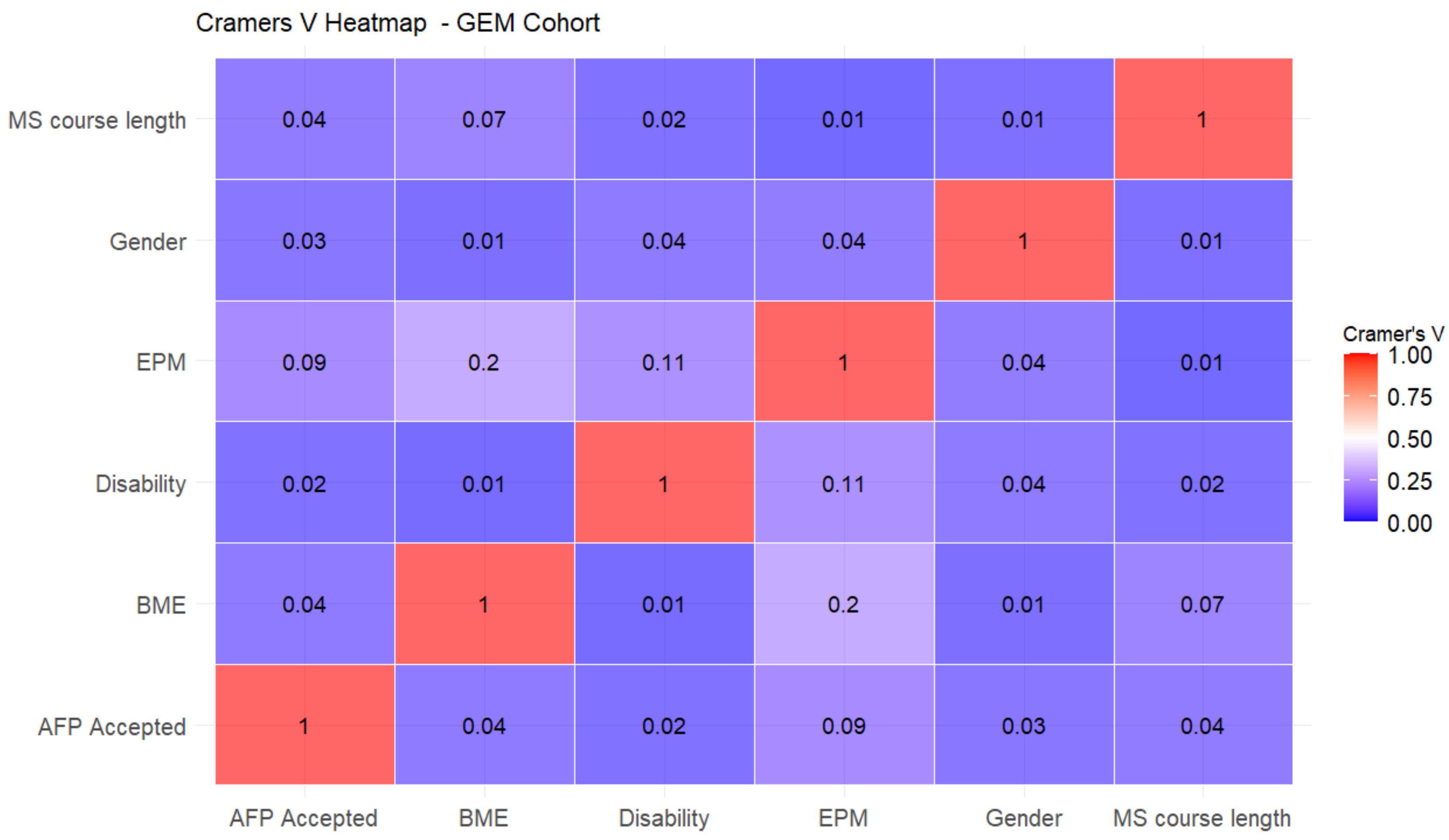


**Supplementary Figure 2: Cramer’s V showing the association of demographic variables in the GE cohort.**

**Supplementary Table 1. Proportions of UG full cohort intercalating, applying to AFP and being accepted to AFP by sociodemographic and education factors**. Significant associations ($X^2$ test, $p < 0.05$) are shown in bold.

| Group | Group value | SE | | | GE | |
|---|---|---|---|---|---|---|
| | | Intercalation N (%) | AFP - applied N(%) | AFP - accepted N (%) | AFP - applied N(%) | AFP - accepted N (%) |
| **Gender** | Female | **8680 (61.1)** | **2635 (18.6)** | 935 (35.5) | **830 (20)** | 235 (28.4) |
| | Male | **7490 (67.5)** | **2765 (25.0)** | 985 (35.6) | **770 (24.4)** | 220 (28.9) |
| **Ethnicity** | Asian/Asian British | **4265 (66.5)** | **1630 (25.4)** | 585 (35.9) | | |
| | Black/Black British | **580 (67.3)** | **200 (23.2)** | 65 (32.5) | | |
| | Mixed | **840 (71.9)** | **265 (22.8)** | 95 (36.1) | | |
| | Other | **440 (64.9)** | **210 (30.5)** | 60 (28.4) | | |
| | White | **10045 (62.1)** | **3100 (19.2)** | 1115 (36) | | |
| **Ethnicity Group** | Ethnicity minority | **6125 (67.2)** | **2305 (25.3)** | 805 (34.9) | 475 (22.7) | **70 (14.7)** |
| | White | **10045 (62.1)** | **3100 (19.2)** | 1115 (36.0) | 1125 (21.6) | **250 (22.1)** |
| **Disability** | Declared | **2035 (60.1)** | 755 (22.2) | 255 (34.1) | 350 (23.2) | **85 (24.3)** |
| | None declared | **14135 (64.5)** | 4650 (21.2) | 1665 (35.8) | 1250 (21.6) | **375 (30)** |
| **IMD** | Low | **2835 (58.5)** | 1040 (21.4) | 345 (33.3) | | |
| | High | **13335 (65.2)** | 4365 (21.4) | 1575 (36.1) | | |
| **School Type** | Privately funded | **5050 (70.9)** | **1775 (24.9)** | **650 (36.6)** | | |
| | State-funded | **11125 (61.2)** | **3630 (20.0)** | **1270 (35.0)** | | |
| **Course Length** | 6-year course | **6015 (92.3)** | **1990 (30.5)** | 705 (35.4) | | |
| | 5-year course | **10160 (54.1)** | **3415 (18.2)** | 1215 (35.6) | | |
| **Medical School Founded** | Pre-1992 | **14775 (67.4)** | 4665 (21.3) | 1640 (35.2) | 1380 (22.3) | 400 (29.2) |
| | Post-1992 | **1395 (41.6)** | 735 (22.0) | 280 (38.0) | 220 (19.7) | 55 (25.3) |
| **Medical School Performance Rank** | Top half | | **3655 (27.7)** | **1525 (41.7)** | **1035 (25.4)** | **335 (32.3)** |
| | Bottom half | | **1750 (14.5)** | **400 (22.9)** | **565 (17.5)** | **125 (21.8)** |

| | | | | | |
|---|---|---|---|---|---|
| **Publication** | Yes | [redacted] | **1585 (47.7)** | **770 (48.6)** | [redacted] |
| | No | [redacted] | **3820 (17.4)** | **1150 (30.1)** | [redacted] |
| **Intercalation** | Yes | [redacted] | **4870 (30.1)** | **1825 (37.5)** | [redacted] |
| | No | [redacted] | **535 (5.9)** | **95 (17.8)** | [redacted] |

**Supplementary Table 2. Proportions applying to and being accepted to a pre-doctoral fellowship (PF) by group, sociodemographic and education factors.** Significant associations ($X^2$ test, $p < 0.05$) are shown in bold.

| Group | Group value | SE | | GE | |
|---|---|---|---|---|---|
| | | **PF - applied N (%)** | **PF - accepted N (%)** | **PF - applied N (%)** | **PF - accepted N (%)** |
| **Gender** | Female | **495 (6.4)** | 185 (37.3) | 135 (5.5) | 30 (23.3) |
| | Male | **555 (8.9)** | 210 (37.6) | 120 (6.6) | 30 (26.7) |
| **Ethnicity Group** | Ethnic Minority | **440 (8.5)** | 135 (30.7) | 75 (6.0) | 15 (Supp) |
| | White | **605 (6.9)** | 360 (59.5) | 180 (6.0) | 50 (27.8) |
| **Disability** | Declared | 130 (7.6) | 50 (38.5) | 55 (6.3) | 15 (Supp) |
| | None declared | 920 (7.5) | 345 (37.3) | 200 (5.9) | 45 (23.0) |
| **IMD** | Low | 185 (6.8) | 55 (31.0) | | |
| | High | 865 (7.7) | 335 (38.8) | | |
| **School Type** | Privately funded | **360 (8.9)** | **155 (43)** | | |
| | State-funded | **690 (6.9)** | **240 (34.6)** | | |
| **Course Length** | 6-year course | 425 (11.4) | 155 (36.6) | | |
| | 5-year course | 625 (6.1) | 235 (37.6) | | |
| **Medical School Founded** | Pre-1992 | **930 (7.7)** | 340 (36.6) | **220 (12.2)** | 50 (23.5) |
| | Post-1992 | **120 (6.3)** | 55 (44.5) | **30 (1.3)** | 10 (Supp) |
| **Medical School Performance Rank** | Top half | **700 (9.6)** | **305 (43.6)** | **160 (6.7)** | 45 (26.5) |
| | Bottom half | **350 (5.2)** | **90 (25.8)** | **90 (5.0)** | 20 (Supp) |
| **Publication** | Yes | **430 (19.3)** | **200 (46.5)** | | |
| | No | **620 (5.3)** | **195 (31.5)** | | |
| **Intercalation** | Yes | **950 (10.7)** | **370 (38.9)** | | |
| | No | **100 (1.9)** | **20 (22.4)** | | |
| **AFP accepted** | Yes | **455 (38.8)** | **245 (53.8)** | **90 (29.9)** | 20 (Supp) |
| | No | **595 (4.6)** | **160 (26.7)** | **165 (4.1)** | 45 (26.4) |

**Supplementary Table 3. Odds ratios and 95% confidence intervals from the logistic regression model comparing graduate-entry and standard-entry applicants applying to the Academic Foundation Programme (AFP) across available demographic characteristics.** The table includes both main effects and interaction terms. Main effects (e.g., gender, ethnicity, disability status, EPM score, and medical school type) compare each category to its reference group. The 'Graduate on entry' comparison treats standard-entry as the reference group with an odds ratio of 1.00. Interaction terms (shaded) reflect the adjusted odds for graduate-entry within specific subgroups, relative to their standard-entry counterparts.

| Reference | Comparison | Odds Ratio (OR) | Lower CI | Upper CI | p value |
|---|---|---|---|---|---|
| **(Intercept)** | (Intercept) | 0.26 | 0.25 | 0.28 | < 0.001 |
| **Gender: Female** | Gender: Male | 1.55 | 1.46 | 1.65 | < 0.001 |
| **Graduate on entry: No** | Graduate on entry: Yes | 1.09 | 0.98 | 1.22 | 0.12 |
| **Ethnicity Group: White** | Ethnicity Group: BME | 1.64 | 1.54 | 1.75 | < 0.001 |
| **Disability: No declared disability** | Disability: Declared disability | 1.24 | 1.13 | 1.36 | < 0.001 |
| **EPM: High** | EPM: Low | 0.39 | 0.36 | 0.42 | < 0.001 |
| **MS Founded: Pre-1992** | MS Founded: Post-1992 | 1.04 | 0.95 | 1.14 | 0.39 |
| **Gender: Female, Graduate on entry: No** | Gender: Male, Graduate on entry: Yes | 0.86 | 0.75 | 0.97 | <0.05 |
| **Ethnicity Group: White, Graduate on entry: No** | Ethnicity Group: BME, Graduate on entry: Yes | 0.74 | 0.64 | 0.85 | < 0.001 |
| **Disability: No declared disability, Graduate on entry: No** | Disability: Declared disability, Graduate on entry: Yes | 0.96 | 0.81 | 1.13 | 0.59 |
| **EPM: High, Graduate on entry: No** | EPM: Low , Graduate on entry: Yes | 1.50 | 1.31 | 1.71 | < 0.001 |
| **MS Founded: Pre-1992, Graduate on entry: No** | MS Founded: Post-1992, Graduate on entry: Yes | 0.83 | 0.69 | 0.99 | <0.05 |

**Supplementary Table 4. Odds ratios and 95% confidence intervals from the logistic regression model comparing graduate-entry and standard-entry applicants being accepted to the Academic Foundation Programme (AFP) across available demographic characteristics**. The table includes both main effects and interaction terms. Main effects (e.g., gender, ethnicity, disability status, EPM score, and medical school type) compare each category to its reference group. The 'Graduate on entry' comparison treats standard-entry as the reference group with an odds ratio of 1.00. Interaction terms (shaded) reflect the adjusted odds for graduate-entry within specific subgroups, relative to their standard-entry counterparts.

| Reference | Comparison | Odds Ratio (OR) | Lower CI | Upper CI | p value |
|---|---|---|---|---|---|
| **(Intercept)** | (Intercept) | 0.09 | 0.08 | 0.10 | < 0.001 |
| **Gender: Female** | Gender: Male | 1.50 | 1.36 | 1.65 | < 0.001 |
| **Graduate on entry: No** | Graduate on entry: Yes | 0.96 | 0.80 | 1.15 | 0.69 |
| **Ethnicity Group: White** | Ethnicity Group: BME | 1.58 | 1.44 | 1.75 | < 0.001 |
| **Disability: No declared disability** | Disability: Declared disability | 1.21 | 1.05 | 1.39 | < 0.05 |
| **EPM: High** | EPM: Low | 0.23 | 0.21 | 0.26 | < 0.001 |
| **MS Founded: Pre-1992** | MS Founded: Post-1992 | 1.12 | 0.98 | 1.28 | 0.09 |
| **Gender: Female, Graduate on entry: No** | Gender: Male, Graduate on entry: Yes | 0.86 | 0.70 | 1.07 | 0.18 |
| **Ethnicity Group: White, Graduate on entry: No** | Ethnicity Group: BME, Graduate on entry: Yes | 0.52 | 0.41 | 0.67 | < 0.001 |
| **Disability: No declared disability, Graduate on entry: No** | Disability: Declared disability, Graduate on entry: Yes | 0.77 | 0.58 | 1.02 | 0.07 |
| **EPM: High, Graduate on entry: No** | EPM: Low , Graduate on entry: Yes | 1.95 | 1.52 | 2.49 | < 0.001 |
| **MS Founded: Pre-1992, Graduate on entry: No** | MS Founded: Post-1992, Graduate on entry: Yes | 0.70 | 0.51 | 0.96 | < 0.05 |

**Supplementary Table 5. Odds ratios and 95% confidence intervals from the logistic regression model comparing graduate-entry and standard-entry applicants applying to an pre-doctoral fellowship across available demographic characteristics.** The table includes both main effects and interaction terms. Main effects (e.g., gender, ethnicity, disability status, EPM score, and medical school type) compare each category to its reference group. The ‘Graduate on entry’ comparison treats standard-entry as the reference group with an odds ratio of 1.00. Interaction terms (e.g., Gender: Male, Graduate on entry: Yes) reflect the adjusted odds for graduate-entry within specific subgroups, relative to their standard-entry counterparts. For example, the interaction term for ‘Gender: Male, Graduate on entry: Yes’ compares graduate-entry males to standard-entry females.

| Reference | Comparison | Odds Ratio (OR) | Lower CI | Upper CI | p value |
|---|---|---|---|---|---|
| **(Intercept)** | (Intercept) | 0.05 | 0.04 | 0.05 | < 0.001 |
| **Gender: Female** | Gender: Male | 1.34 | 1.17 | 1.53 | < 0.001 |
| **Graduate on entry: No** | Graduate on entry: Yes | 0.88 | 0.66 | 1.17 | 0.38 |
| **Ethnicity Group: White** | Ethnicity Group: BME | 1.22 | 1.06 | 1.41 | < 0.05 |
| **Disability: No declared disability** | Disability: Declared disability | 1.11 | 0.90 | 1.36 | 0.32 |
| **EPM: High** | EPM: Low | 0.75 | 0.65 | 0.87 | < 0.001 |
| **MS Founded: Pre-1992** | MS Founded: Post-1992 | 0.74 | 0.60 | 0.91 | < 0.05 |
| **AFP acceptance: No** | AFP acceptance: Yes | 11.89 | 10.24 | 13.81 | < 0.001 |
| **Gender: Female, Graduate on entry: No** | Gender: Male, Graduate on entry: Yes | 0.87 | 0.65 | 1.18 | 0.37 |
| **Ethnicity Group: White, Graduate on entry: No** | Ethnicity Group: BME, Graduate on entry: Yes | 0.96 | 0.69 | 1.33 | 0.80 |
| **Disability: No declared disability, Graduate on entry: No** | Disability: Declared disability, Graduate on entry: Yes | 1.09 | 0.73 | 1.59 | 0.68 |
| **EPM: High, Graduate on entry: No** | EPM: Low , Graduate on entry: Yes | 1.14 | 0.83 | 1.57 | 0.42 |
| **MS Founded: Pre-1992, Graduate on entry: No** | MS Founded: Post-1992, Graduate on entry: Yes | 1.16 | 0.73 | 1.8 | 0.516 |
| **AFP acceptance: No, Graduate on entry: No** | AFP acceptance: Yes, Graduate on entry: Yes | 0.81 | 0.58 | 1.13 | 0.226 |
